# Study On the Changes of Pattern Reversal Visual Evoked Potential In PSCI Patients

**DOI:** 10.1101/2025.06.11.25329465

**Authors:** Min Gao, Xin-yu Miao, Lu Huang, Xin-yi Li, Wei-bo Tian, Shu-yue Yu, Xiao-qin Duan

## Abstract

**Objective:** To study the pattern reversal visual evoked potential (PRVEP) in patients with post stroke cognitive impairment (PSCI) and to provide an objective and accurate basis for the neurophysiological assessment of cognitive function in stroke patients.

**Methods:** First stroke patients admitted to the department of Rehabilitation Medicine of the Second Hospital of Jilin University between 01-10-2022 and 31-10-2023 were enrolled as subjects according to the inclusion criteria, exclusion criteria, shedding and exclusion criteria. They were screened for whole cognitive function using the Montreal Cognitive Assessment (MoCA), and were divided into a post-stroke cognitive impairment (PSCI) group (n=44) and a control group (post-stroke patients with normal cognitive function, n=25). The PSCI group was divided into three group including mild, moderate and severe PSCI patients according to the severity of their cognitive impairment. Clinical data of the above patients were collected, PRVEP examination was performed and index of P100 waves were calculated for both eyes. SPSS 25.0 statistical analysis was used to analyze data of patients.

**Results:** 69 first-stroke patients completed the whole experiment, including 25 cases in control group, 14 cases in mild PSCI group, 14 cases in moderate PSCI group, and 16 cases in severe PSCI group. In patients with detectable VEP, the difference in latent period of P100 in both eyes of the PSCI group (n=20) was greater than that of the control group (n=19) (*P*<0.01), and the amplitude ratio of P100 waves on both sides in PSCI group was higher than that of the control group (*P*<0.05). Compared with PSCI patients with left-sided hemiplegia, PSCI patients with right-sided hemiplegia had a longer P100 latency in the right eye (*P*<0.05); Compared with PSCI patients without brainstem involvement, PSCI patients with brainstem involvement had an increased P100 latency in both eyes (*P*<0.01); Pearson correlation analysis showed that the binocular amplitude ratio in the VEP parameters in the PSCI group was correlated with the MoCA score (*r*=-0.624, *P*<0.01). The ROC curve showed that the difference in the latent period of P100 in both eyes and the ratio of P100 amplitude in both eyes had certain predictive value for the diagnosis of PSCI (AUC=0.875, 0.842; *P*<0.05).

**Conclusion:** PRVEP examination and VEP parameters can help to distinguish stroke patients with or without cognitive impairment. In first-stroke patients with detectable VEP, the difference in binocular P100 latency and the ratio of binocular P100 amplitude have certain predictive value for diagnosis of PSCI, which is worthy of further study and application.

## Background

The Global Burden of Disease Study (GBD) underscores that stroke stands as the foremost cause of mortality and disability among Chinese residents, with China leading globally in stroke prevalence[1]. Post stroke cognitive impairment (PSCI) emerges as the most prevalent complication following stroke, delineated by cognitive functional impairments persisting up to six months post-stroke[1]. Surveys reveal that as high as 43.6% of stroke patients manifest cognitive impairment[4]. Research indicates that individuals afflicted with stroke are at least 5 to 8 times more likely to develop cognitive impairment[5]. Cognitive dysfunction commonly manifests in the early stages post-stroke, yet its onset remains insidious. Not only does it detrimentally impact functions such as attention, executive function, memory, language, and visuospatial abilities, but it also hampers the overall rehabilitation trajectory of patients, significantly augmenting rates of disability and mortality[5]. The mechanisms underlying post-stroke cognitive impairment remain elusive, although vascular cognitive impairment (VCI) or Alzheimer’s disease (AD) precipitated by stroke are posited as potential etiologies. Certain studies suggest their synergistic involvement in post-stroke cognitive impairment. With the burgeoning burden of stroke, PSCI has emerged as an increasingly pressing public healthcare challenge, underscoring the paramount importance of early screening and diagnosis for PSCI.

Due to the lack of distinctive biological and radiological markers**Error! Reference source not found**., the clinical diagnosis of Post-Stroke Cognitive Impairment (PSCI) heavily relies on cognitive assessments such as the Mini-Mental State Examination (MMSE), Montreal Cognitive Assessment (MoCA), Addenbrooke’s Cognitive Examination-Revised (ACE-R), and Wechsler Adult Intelligence Scale (WAIS). However, assessments based on neuropsychological evaluation scales often exhibit subjectivity and uncertainty[8], susceptible to influences such as age and cultural backgroundF[9]. Consequently, neuropsychological examinations alone are insufficient for reliably diagnosing and prognosticating PSCI. Hence, the exploration of objective and effective methods for assessing and diagnosing post-stroke cognitive impairment assumes paramount importance.

In recent years, numerous domestic and international studies have proposed Visual Evoked Potentials (VEP) as a robust adjunctive measure for assessing stroke. VEPs entail electroencephalographic responses evoked in the occipital cortex by visual stimuli received by the retina, ultimately recorded on the scalp[10]. The most commonly utilized stimulus in clinical practice for Pattern Reversal Visual Evoked Potentials (PRVEP) is the checkerboard reversal stimulus, which induces retinal stimulation through the reversal of checkerboard squares at different temporal and spatial frequencies. This elicits neuroelectric responses in the occipital cortex, with the latency of the P100 component reflecting the speed of signal transmission from retinal ganglion cell axons to the visual cortex and the visual system’s capacity to process spatial frequency information. Furthermore, the amplitude of the P100 wave partly mirrors the physiological state of the ganglion cells. Given that VEP serves as a means to detect functional alterations and abnormalities, it can detect anomalies clinically even in the absence of evident structural changes in the early stages of stroke, thereby serving as a potential reference for early diagnosis to a certain extent. Regrettably, there is currently a dearth of literature both domestically and internationally regarding the application of PRVEP assessment in patients with PSCI. Hence, this study aims to investigate the distinctive PRVEP variations in individuals with PSCI and those with varying degrees of cognitive dysfunction. By doing so, it seeks to furnish a foundation of objective and precise neurophysiological evaluation for cognitive function in stroke patients.

## Material and methods

### 1 Study design

A cross-sectional survey was occurred between 01-10-2022 and 31-10-2023 at the Rehabilitation Department of the Second Hospital of Jilin University to assess cognitive function of first-stroke patients with PRVEP. The method used in the study was reviewed and approved by the Ethics Review Board of the Second Hospital of Jilin University (No 2021-104) and we confirm that all methods were performed in accordance with the relevant guidelines and regulations. Patients provided informed consent and signed informed consent forms.

### 2 Participants

#### 2.1 Inclusion Criteria

First-stroke patients, with brain CT or MRI indicating stroke. Conformance with the diagnostic criteria for Post-Stroke Cognitive Impairment (PSCI) outlined in the 2017 “Expert Consensus on the Management of Post-Stroke Cognitive Impairment”. MoCA score below 23 points, attributable to the stroke occurring within 3 to 12 months, if the patient’s educational level is ≤12 years, 1 point is added to the test result.

#### 2.2 Exclusion Criteria

History of brain diseases other than cerebrovascular diseases, such as traumatic brain injury, brain infections, subdural hematoma, etc. Patients with cognitive impairment caused by Parkinson’s disease, Alzheimer’s disease, Lewy body dementia, etc. Patients with consciousness disorders, neurodegenerative diseases, or severe neurological impairments. Inability to complete the assessment due to auditory, visual, or language impairments. Personal or family history of epilepsy or psychiatric disorders. Presence of other diseases affecting visual function, such as glaucoma, cataracts, etc. Failure to sign the informed consent form.

#### 2.3 Dropout and Exclusion Criteria

Significant interference or unclear waveform in VEP recording. Missing data in the collection of general information and observation indicators for patients.

## 3. Methods

The specific research methods, experimental procedures and key techniques are outlined below.

### 3.1 Clinical material

Demographic data, including gender, age, years of education, alcohol consumption, smoking history, site of stroke occurrence, and medical history (e.g., diabetes, hypertension, coronary artery disease, history of ischemic or hemorrhagic stroke), were collected. Additionally, all enrolled patients underwent MRI within one week of admission to determine the lesion location, whether the lesion was singular or multiple, and whether the lesion was located in the left hemisphere, right hemisphere, or both hemispheres of the brain.

### 3.2 Cognitive function assessment

MoCA served as a rapid screening tool for cognitive function, comprising 13 items across seven domains: visuospatial abilities, executive function, naming, memory, attention, language, and abstraction. Scores range from 0 to 30, with higher scores indicating better cognitive function. An additional point is added to the total score if the patient’s education level is ≤12 years**Error! Reference source not found**.. A MoCA score <23 was used as the cutoff for cognitive impairment**Error! Reference source not found**., with scores of 18–22 indicating mild impairment (mild PSCI group), 10–17 indicating moderate impairment (moderate PSCI group), and <10 indicating severe impairment (severe PSCI group).

### 3.3 PRVEP Examination

The procedure and recording were conducted using the Nicolet Viking Quest electromyography/evoked potential instrument within a shielded room. A full-field 16×16 black-and-white checkerboard pattern reversal stimuli were employed to stimulate the left and right eyes, with evoked potentials recorded from the visual cortex. The recording electrode (Oz) was placed at the occiput, the reference electrode was placed beneath both ears, and the ground electrode could be placed on the hand. Detection parameters: Stimulation frequency of 2 Hz, spatial frequency of 15 Hz, checkerboard contrast of 97%, and averaging over 100 stimuli or more. Computer-averaged data were obtained, and after waveform stabilization, N75 (ms), P100 (ms), N145 (ms) latency, and P100 peak amplitude (μV) were marked and analyzed.

## 4. Statistical Analysis

Data analysis was conducted using SPSS version 25.0 statistical software. Continuous variables are presented as mean (± standard deviation) and compared between groups using the Student’s t-test for normally distributed data. Categorical variables are expressed as percentages (%) and compared using the chi-square test. Pearson correlation analysis was performed to assess the correlation between VEP parameters and MoCA scores in PSCI patients. One-way ANOVA was used to analyze differences among PSCI patient groups of different severities. Receiver operating characteristic (ROC) curves were plotted, and the area under the curve (AUC) was calculated to evaluate the predictive value of pattern reversal visual evoked potential (PRVEP) examination for diagnosing PSCI patients. A significance level of *P* < 0.05 was considered statistically significant.

## Results

### 4.1 General information comparison

Among the 73 selected participants, information collection was omitted for 2 individuals, and VEP wave interference was significant for 2 others. Eventually, 69 individuals completed the entire experimental procedure. Among them, there were 25 cases in the post-stroke patients with normal cognitive function (control group), 14 cases in the mild PSCI group, 14 cases in the moderate PSCI group, and 16 cases in the severe PSCI group. General information of each group was as follows (Table 1).

**Table 1.** Comparison of General Information 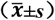.

| Characteristics |  | Control<br>(n=25) | Mild PSCI<br>(n=14) | Moderate PSCI<br>(n=14) | Severe PSCI<br>(n=16) |
| --- | --- | --- | --- | --- | --- |
| Gender (n) | Male | 13 | 6 | 5 | 9 |
|  | Female | 12 | 8 | 9 | 7 |
| Age (years) | | 63.29±6.21 | 61.72±10.08 | 67.57±8.73 | 62.38±9.26 |
| MoCA Score | | 26.29±1.11 | 19.43±1.40 | 13.71±3.15 | 4.00±2.62 |

### 4.2 PRVEP among PSCI Patients

#### 4.2.1 The occurrence rate of VEP

There was no significant difference in the incidence of VEP between the PSCI patient group and the control group (45.5% vs 76.0, *P*>0.05) (Table 2).

**Table 2:** Occurrence Rates of VEP in the PSCI group and the Control group.

|  | Control group | PSCI group |
| --- | --- | --- |
| Number of Patients | 25 | 44 |
| Number of VEP | 19 | 20 |
| Occurrence Rate of VEP (%) | 76.0 | 45.5 |

\*Note: Compared to the control group, \* $P<0.05$ , \*\* $P<0.01$ .

#### 4.2.2 Changes in VEP parameters

VEP parameters of the PSCI group (n=20) and the control group (n=19) exhibited a normal distribution. There was no significant statistical difference in the latency and amplitude of P100 waves between the PSCI group and the control group (*P*>0.05). However, the difference in latent period of P100 in both eyes of the PSCI group was greater than that of the control group (*P*<0.01), and the amplitude ratio of P100 waves on both sides in PSCI group was higher than that of the control group (*P*<0.05)., as shown in Table 3.

**Table 3.**
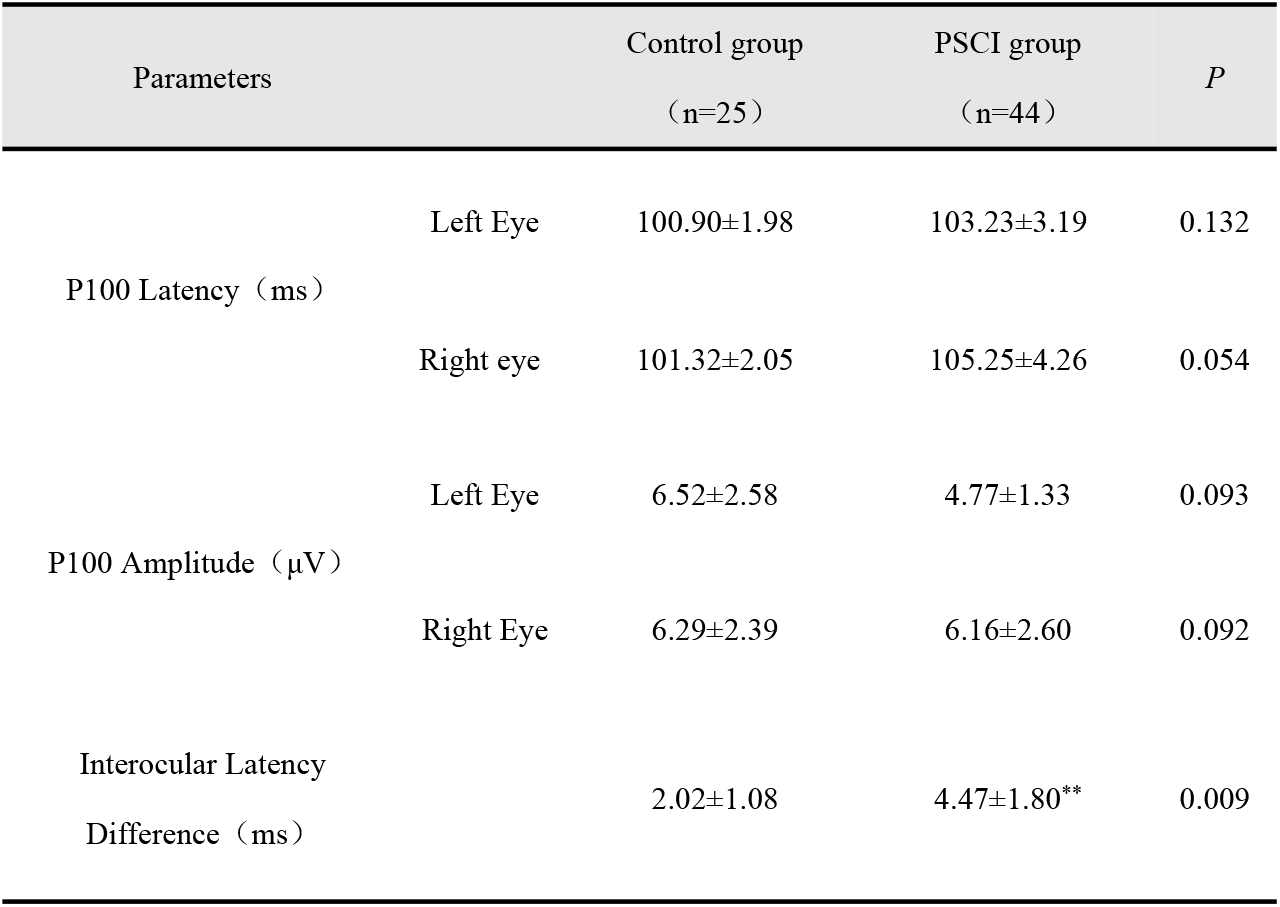
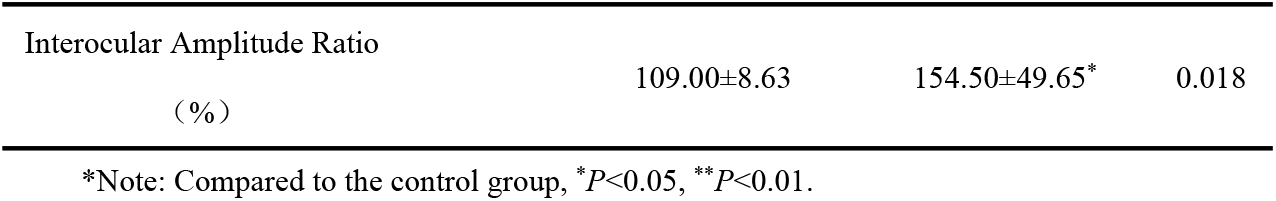
Comparison of VEP parameters between the PSCI group and the Control group 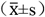.

#### 4.2.3 Comparison of VEP parameters in PSCI patients with different hemiplegic sides

Compared with PSCI patients with left-sides hemiplegia, those with right-sides hemiplegia had a prolonged latency of P100 in the right eye (*P*<0.05), and a decreased amplitude of P100 in both eyes (*P*<0.05, *P*<0.01),. There was no statistically significant difference in latency and amplitude ratio between the two eyes (see Table 4).

**Table 4.** Comparison of VEP parameters in PSCI patients with different hemiplegic sides 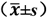.

|  |  | left-sides hemiplegia<br>(n=11) | right-sides hemiplegia<br>(n=9) | *P* |
| --- | --- | --- | --- | --- |
| P100 Latency (ms) | Left Eye | 100.86±3.35 | 103.80±3.11 | 0.188 |
|  | Right Eye | 99.36±2.69 | 105.4±4.98* | 0.044 |
| P100 Amplitude (μV) | Left Eye | 6.81±1.51 | 4.28±0.84* | 0.014 |
|  | Right Eye | 8.85±1.88 | 4.52±1.54** | 0.004 |
| Interocular Latency<br>Difference (ms) |  | 2.60±1.97 | 3.55±2.20 | 0.493 |
| Interocular Amplitude<br>Ratio (%) |  | 141.20±54.28 | 116.20±11.10 | 0.343 |

\*Note: Compared to PSCI patients with left-sides hemiplegia, \* $P<0.05$ , \*\* $P<0.01$ .

#### 4.2.4 Comparison of VEP parameters in PSCI patients with different stroke sites

Among PSCI patients with detectable VEP (n=20), the stroke sites are shown in Table 5. Compared with patients without brainstem involvement, PSCI patients with brainstem involvement showed an increased P100 latency in both eyes (*P*<0.01). However, there was no statistically significant difference in VEP parameters between PSCI patients with involvement of the parietal lobe, temporal lobe, basal ganglia, corona radiata, cerebellum, thalamus, frontal lobe, and occipital lobe and those without above sites involved (*P*>0.05)

**Table 5.** Comparison of VEP parameters in PSCI patients with different stroke sites 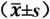.

| Stroke sites |  | Interocular latency difference of P100<br>(ms) | P |
| --- | --- | --- | --- |
| Brainstem | Yes (n=8) | 4.94±1.51** | 0.007 |
|  | No (n=12) | 1.83±1.21 |  |
| Parietal Lobe | Yes (n=6) | 1.50±0.43 | 0.112 |
|  | No (n=14) | 3.75±2.09 |  |
| Temporal Lobe | Yes (n=6) | 1.50±0.43 | 0.112 |
|  | No (n=14) | 3.75±2.09 |  |
| Subarachnoid Space | Yes (n=0) | —— | —— |
|  | No (n=20) | 3.08±2.03 |  |
| Basal Ganglia | Yes (n=8) | 2.13±1.45 | 0.250 |
|  | No (n=12) | 3.71±2.23 |  |
| Corona Radiata | Yes (n=6) | 1.58±1.18 | 0.136 |
|  | No (n=14) | 3.71±2.04 |  |
| Cerebellum | Yes (n=2) | 1.94±1.25 | 0.153 |
|  | No (n=18) | 3.39±1.88 |  |
| Thalamus | Yes (n=2) | 3.75±1.89 | 0.784 |
|  | No (n=18) | 3.00±2.14 |  |
| Frontal Lobe | Yes (n=2) | 2.00±1.45 | 0.608 |
|  | No (n=18) | 3.19±2.12 |  |
| Occipital Lobe | Yes (n=8) | 2.06±1.18 | 0.217 |
|  | No (n=12) | 3.75±2.29 |  |

\*Note: Compared to PSCI patients without stroke occurring in the site, \* $P<0.05$ , \*\* $P<0.01$ .

#### 4.2.5 Correlation analysis between VEP parameters and MoCA scores in PSCI group

Using Pearson correlation analysis, it was observed that there was a correlation between the interocular amplitude ratio of VEP parameters and MoCA scores in the PSCI group (r=-0.624, *P*<0.01), as shown in Table 6.

**Table 6.** Pearson correlation analysis between VEP parameters and MoCA scores in PSCI group.

|  | P100<br>Latency<br>(Left) | P100<br>Amplitude<br>(Left) | P100<br>Latency<br>(Right) | P100<br>Amplitude<br>(Right) | Interocular<br>Latency<br>Difference | Interocular<br>Amplitude<br>Ratio |
| --- | --- | --- | --- | --- | --- | --- |
| MoCA Score<br>(r) | -0.150 | 0.331 | -0.488 | 0.101 | -0.458 | -0.624** |
| *P* | 0.580 | 0.211 | 0.055 | 0.101 | 0.075 | 0.010 |

\*Note: \* $P<0.05$ , \*\* $P<0.01$ .

### 4.3 Variations in PRVEP among patients with different degree of PSCI

#### 4.3.1 Occurrence rate of VEP among different PSCI groups

There was no significant difference in the occurrence rate of VEP among different PSCI groups (*P*>0.05, Table 7).

**Table 7.** Comparison of Occurrence rate of VEP among different PSCI group.

|  | Present | Absent |
| --- | --- | --- |
| Mild PSCI | 6/14 (42.86%) | 8/14 (57.14%) |
| Moderate PSCI | 6/14 (42.86%) | 8/14 (57.14%) |
| Severe PSCI | 8/16 (50.0%) | 8/16 (50.0%) |

\*Note: \* $P<0.05$ , \*\* $P<0.01$ .

#### 4.3.2 Comparison of VEP parameters in different groups of PSCI patients

The results of the analysis of ANOVA showed that there was no significant difference in VEP parameters among patients in different groups of PSCI (*P*>0.05), as shown in Table 8.

**Table 8.**
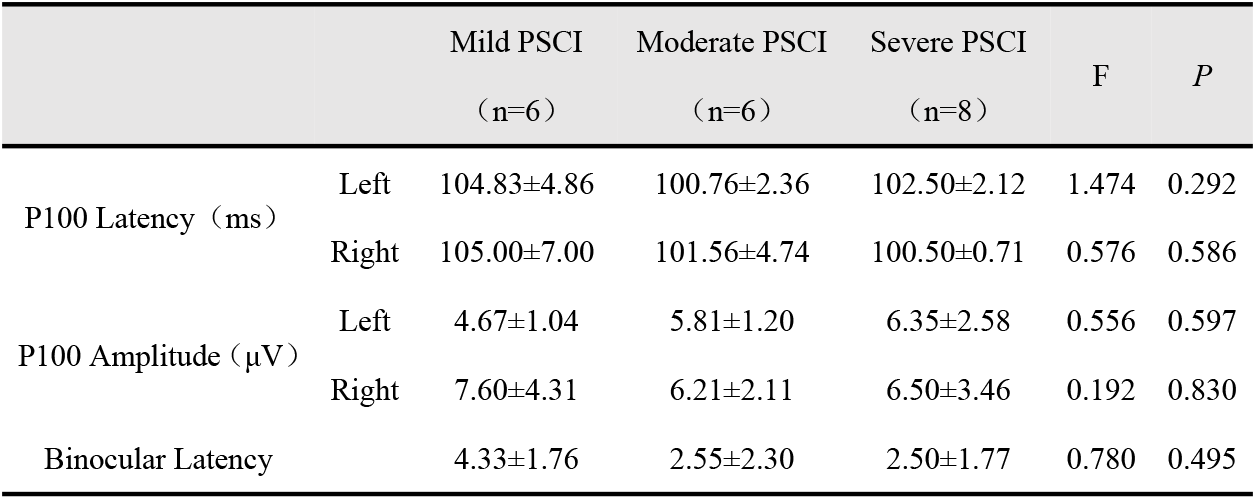
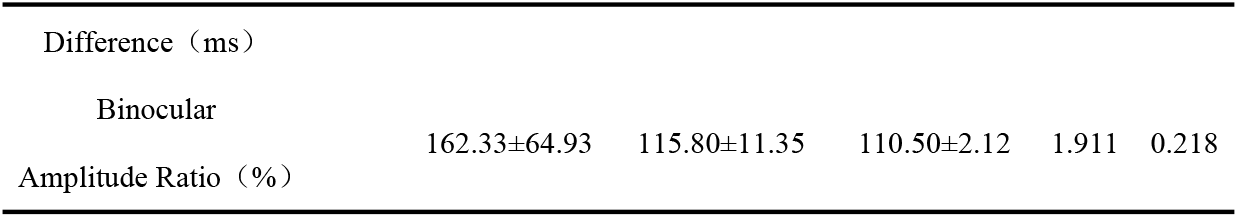
Comparison of VEP parameters in different groups of PSCI patient 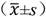.

### 4.4 Diagnostic value of PRVEP changes on PSCI

In the cohort of first-time stroke patients with detectable VEP, ROC curves were constructed using PSCI patients as positive samples and post-stroke cognitively normal patients as negative samples. The results showed that the difference in the latent period of P100 in both eyes and the ratio of P100 amplitude in both eyes had certain predictive value for the diagnosis of PSCI (AUC=0.875, 0.842; *P*<0.05), with thresholds of 3.75 ms and 129.5%, respectively, as shown in Table 9 and Figure 1.

**Fig. 1.**
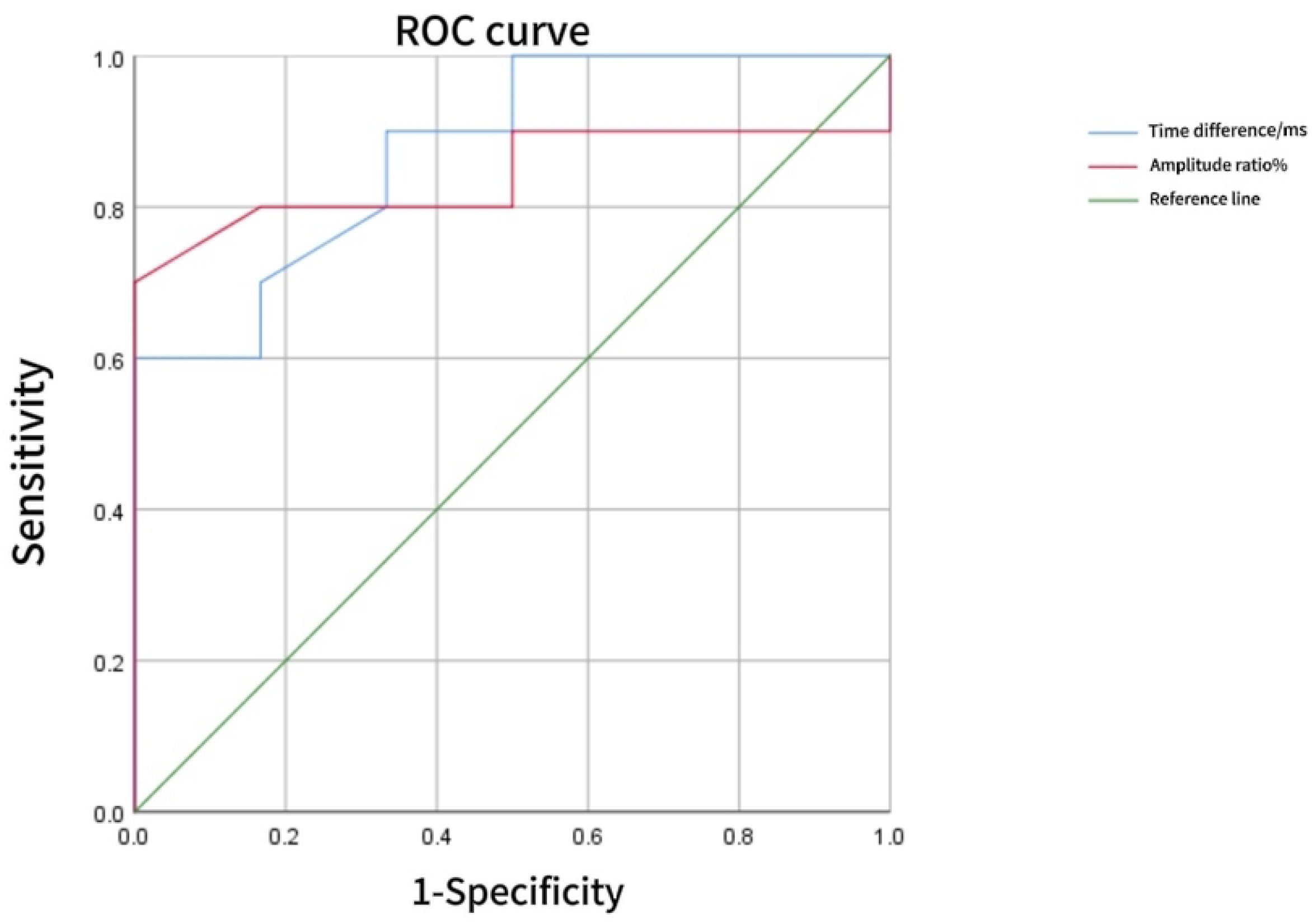

**Table 9.** Diagnostic value of PRVEP changes on PSCI.

|  | AUC | *P* | 95% Confidence Interval |  | Cut-off Value | Sensitivity (%) | Accuracy (%) |
| --- | --- | --- | --- | --- | --- | --- | --- |
| Binocular Latency Difference | 0.875 | 0.015 | 0.702 | 1.000 | 3.75ms | 60 | 87.5 |
| Binocular Amplitude Ratio | 0.842 | 0.026 | 0.634 | 1.000 | 129.50% | 70 | 84.2 |

## Discussion

Research has shown that in the early stages of mild cognitive impairment (MCI) or Alzheimer’s disease (AD), patients exhibit impaired cholinergic function, leading to impaired visual processing and significantly prolonged latency of VEPs [14]. Measuring pattern reversal visual evoked potentials under dual stimulus conditions can provide valuable diagnostic information about the cholinergic system [15]. Although the pathogenesis of PSCI is unclear, vascular cognitive impairment or AD caused by stroke have potential synergistic effects on PSCI.

As an objective electrophysiological indicator, VEP has been widely applied in the clinical diagnosis of neurological and ophthalmological disorders, as well as in monitoring intracranial pressure, diagnosing and treating brain tumors, and intraoperative monitoring during stroke and brain injury. The pathway of VEP transmission includes conduction from the retina to the lateral geniculate nucleus, then through the optic nerve to the occipital visual cortex. The disappearance of waveform may be attributed to poor vision or lack of attention from patients. If these conditions are ruled out, it may indicate abnormalities in the conduction of neural pathways. Absence of responses in one eye may indicate lesions in the affected anterior visual pathway, while prolonged latency mainly reflects demyelination changes in the nerve fibers of the ipsilateral anterior conduction pathway (excluding retinal or optic nerve lesions). Waveform amplitudes mainly reflect changes in the number of axons, with considerable variability among individuals, but primarily implies a reduction in visual sensor input or degeneration of the visual conduction pathway. Any abnormalities affecting the retina, visual pathway, or visual cortex may influence VEP.

Studies have shown that cerebral ischemia caused by local cerebral circulation problems can lead to differences in VEP amplitude and latency between cerebral hemispheres [20]. Abnormal VEP can serve as a reference indicator for evaluating consciousness disorders in stroke patients with subarachnoid hemorrhage. The application value of VEP in stroke patients depends on the location of the lesion. The changes in VEP are more pronounced in posterior circulation stroke, providing valuable auxiliary diagnostic value. Due to the fact that most stroke lesions occur behind the optic chiasm, VEP monitoring has significant observational and evaluative significance in clinical practice. The specific location of the lesion has different effects on the VEP waveform. For stroke in the vertebral basilar artery system, VEP detection is highly abnormal with good reproducibility, indicating potential diagnostic value for PSCI patients with brainstem involvement. The results of this study also suggest that PSCI patients with brainstem involvement have increased P100 latency in both eyes. Furthermore, we find that the difference in the latent period of P100 in both eyes and the ratio of P100 amplitude in both eyes had certain predictive value for the diagnosis of PSCI (*P*<0.05). Therefore, VEP, as a method for detecting functional changes, can provide reference for early diagnosis.

There are several limitations to this study. Firstly, due to the small sample size, our experimental results may not be generalizable and further large-scale clinical studies are needed in the future. Secondly, although visual evoked potential monitoring has clinical utility, it still has limitations such as waveform instability, waveform extraction failure, and significant changes in latency and amplitude of normal values. In addition, this study explored the relationship between VEP parameters and whole cognitive function, and future research can further explore the potential association between visual spatial function, executive function, attention, and other cognitive domains with VEP.

## Conclusion

The PRVEP examination and VEP parameters contribute to distinguishing stroke patients with or without cognitive impairment. In first-stroke patients with detectable VEP, the difference in bilateral P100 latency and the ratio of bilateral P100 amplitude have certain predictive value for diagnosing PSCI, warranting further investigation and application.

## Data Availability

The data underlying the results presented in the study are available from (include the name of the third party and contact information or URL).

## Data Availability Statement

All data generated or analysed during this study are included in this published article.

## Notes

### Competing Interest Statement

The authors have declared no competing interest.

### Funding Statement

The author(s) received no specific funding for this work.

### Author Declarations

The method used in the study was reviewed and approved by the Ethics Review Board of the Second Hospital of Jilin University (No 2021-104).

